# COVID-19 in Japan: What could happen in the future?

**DOI:** 10.1101/2020.02.21.20026070

**Authors:** Nian Shao, Yan Xuan, Hanshuang Pan, Shufen Wang, Weijia Li, Yue Yan, Xingjie Li, Christopher Y. Shen, Xu Chen, Xinyue Luo, Yu Chen, Boxi Xu, Keji Liu, Min Zhong, Xiang Xu, Yu Jiang, Shuai Lu, Guanghong Ding, Jin Cheng, Wenbin Chen

## Abstract

COVID-19 has been impacting on the whole world critically and constantly Since December 2019. We have independently developed a novel statistical time delay dynamic model on the basis of the distribution models from CCDC. Based only on the numbers of confirmed cases in different regions in China, the model can clearly reveal that the containment of the epidemic highly depends on early and effective isolation. We apply the model on the epidemic in Japan and conclude that there could be a rapid outbreak in Japan if no effective quarantine measures are carried out immediately.

## Is it just similar?

The cumulative number of confirmed cases of COVID-19 in Wuhan, China (Jan 25, 2020 –Feb 1, 2020, in blue) and Japan (Feb 14, 2020–Feb 21, 2020, in red) is demonstrated in Figure A very similar linear growth of the epidemic can be observed, at the early stage of the epidemic in Japan and Wuhan, China. We suspect that there might also be similarities in future trend. Therefore, we forecast that the situation in Japan is hardly optimistic, if the current isolation rate of Wuhan is used for Japan. We should concern that there could be a possible severe outbreak in Japan if no strong quarantine strategy is carried out immediately.

## Background

In December 2019, a pneumonia epidemic of unknown cause broke out in Wuhan, Hubei Province, China. The atypical pneumonia was caused by 2019 novel coronavirus, later named as COVID-19 by World Health Organization. As Wuhan is the major transportation hub in central China, the epidemic quickly spread to other major cities in China such as Beijing, Shanghai, Chongqing, Guangzhou and Shenzhen (*1*), and even spread internationally to other countries. As of Feb 20, 2020, there have been 74675 confirmed cases in mainland China and 26 countries have reported imported cases (*20*).

COVID-19 is a group of SARS-like coronaviruses (*2*) and closely related to SARS-CoV and MERS-CoV. All of them are zoonotic viruses and epidemiologically similar. It is reported that human-to-human transmission has occurred among close contacts since the middle of December 2019 on the basis of evidence from early transmission dynamics (*3*). Cases found outside Wuhan also indicate independent self-sustaining human-to-human spread in multiple Chinese major cities (*20*). Thus the risk of epidemic outbreak cannot be neglected outside Wuhan.

The epidemic of COVID-19 has raised intense attention in academia. A few researches focus on the effect of migration and transportation on the spread of epidemic. In (*4*), the authors estimated the probability of transportation of COVID-19 from Wuhan to 369 cities in China before the quarantine of Wuhan and concluded that the expected risk is greater than 50% in 130 (95% CI 89-190) cities and is even greater than 99% in the 4 largest metropolitan areas of China. In (*5*), the authors studied the feasibility of controlling COVID-19 outbreaks by isolation of cases and contacts. Their results suggest that it is very difficult to control the trends of infection under a high value of *R*_0_ and when a large initial number of cases has appeared even if less than 1% of transmission occurred before symptom onset. Similarly, (*1, 6*) also suggested proper transmissibility reduction to avoid the outbreak of epidemic. Case numbers are rising exponentially in multiple cities due to human-to-human transmission. To possibly secure containment of the spread of infection, substantial public health interventions should be considered and carried out immediately. Multiple provinces including Hubei have adopted measures such as metropolitan-wide quarantine and reduction of inter-city mobility. Besides, it is crucial to isolate patients, trace and quarantine all possible contacts as the earliest stage because of the possibility of asymptomatic infections (*7*).

On the other hand, new models on the quantitative analysis of the spread of epidemic have been developed. For instance, (*8*) adopted a novel statistical time delay dynamic system to estimate the basic reproductive number *R*_0_ of COVID-19 based on Wallinga and Lipsitch’s framework (*9*) with distribution of the generation interval of the infection obtained. In (*8*), the growth rate of COVID-19 was estimated to be between 0.30 and 0.32, higher than that estimated by CCDC (Chinese Center for Disease Control and Prevention) (*3*). Meanwhile, the basic reproductive number *R*_0_ of COVID-19 was estimated between 3.25 and 3.4, larger than that of SARS. Paralleled results based on other models could be found in (*1, 5, 10–14*).

In this study, by data fitting, we are able to predict the outbreak trend of COVID-19 in multiple provinces and cities in China. Our model so far is consistent with the observed data in mainland China. We also apply the model to track and predict the spread of COVID-19 in Japan. We observe that the epidemic situation in Japan is quite similar to that in Wuhan, which draws our concerns on a possible severe outbreak in Japan. According to our model, we suggest Japan enhancing public health interventions as soon as possible.

## Model

The data employed in this paper are the cumulative confirmed cases from Jan 16, 2020 to Feb 21, 2020 (for mainland China), and from Jan 20, 2020 to Feb 21, 2020 (for Japan), acquired from the National Health Commission of China (http://www.nhc.gov.cn), and Ministry of Health, Labour, and Welfare, Japan (https://www.mhlw.go.jp/index.html). All the data can be accessed publicly. No other data are used in this paper.

As the data are announced daily, we adopt the following discrete version of the FUDANCCDC model proposed recently in (*8*):

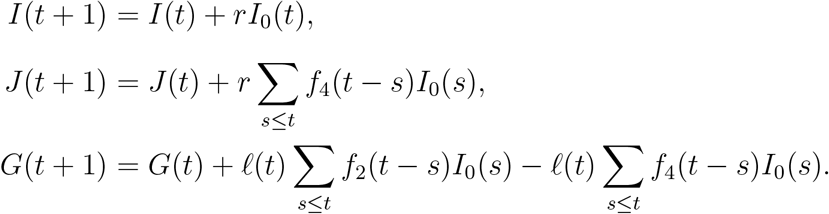

A sketch map of the model is displayed in Figure 2. At day *t*, groups *I*(*t*), *J* (*t*) and *G*(*t*) represent the cumulative number of the infected, the cumulative number of the confirmed (by hospital), and the instant (not cumulative) number of isolated infected not yet confirmed, respectively. The isolated group *G* are infected actually, but not confirmed by the hospital, so these people are not counted by CCDC the infected ones. *I*_0_(*t*) := *I*(*t*) *− J* (*t*) *− G*(*t*) is the number of infected people neither confirmed nor isolated, i.e., the ones who are potentially infectious to healthy ones. The two important parameters *r* and *ℓ* are the growth rate and the isolation rate respectively. *f*_2_(*t*) and *f*_4_(*t*) are the transition probabilities from infection to illness onset, and from infection to hospitalization, respectively. We have reconstructed them from one important paper by CCDC, see Fig 2 in (*3*).

**Figure 1:**
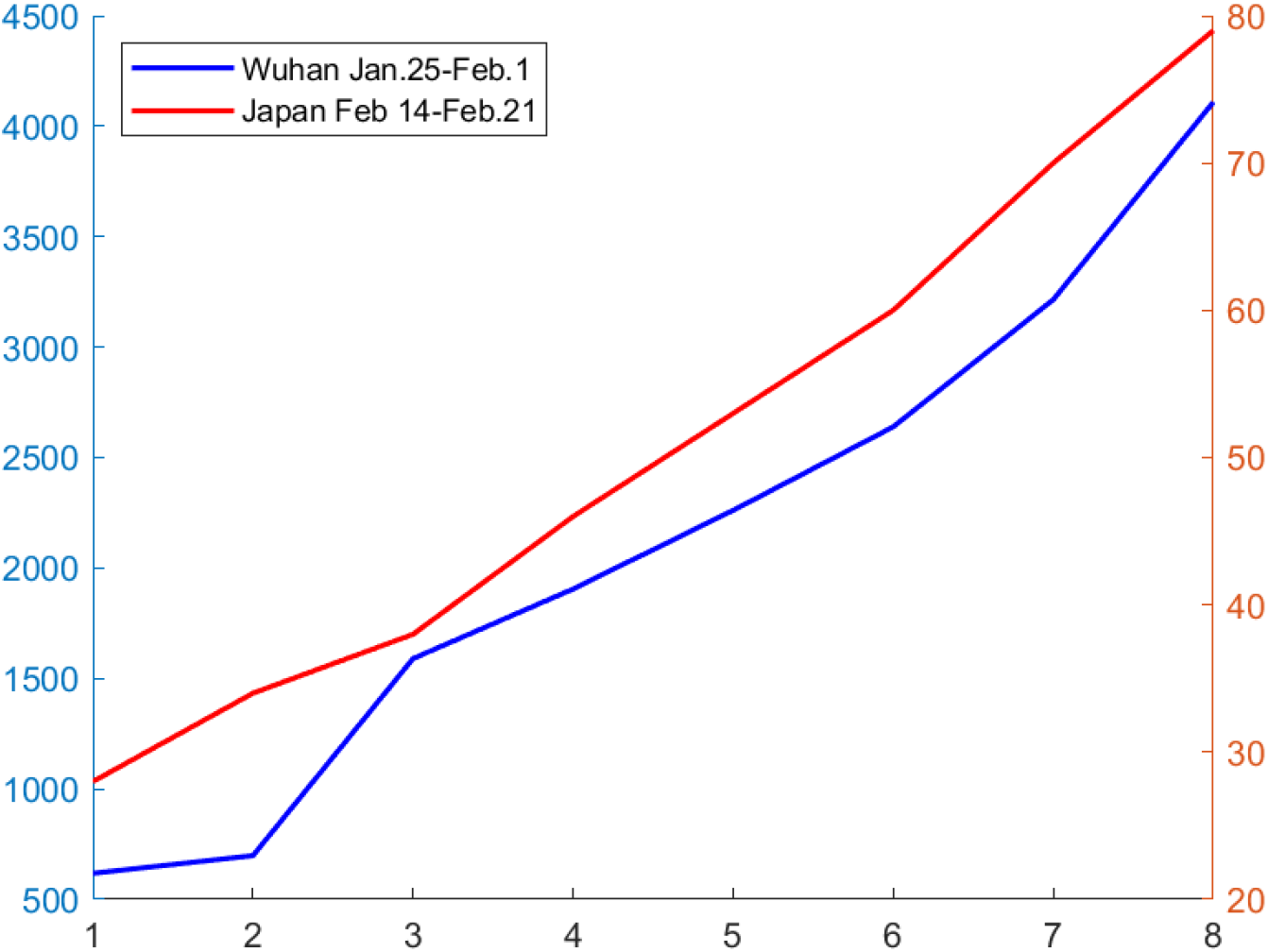
Cumulative number of confirmed cases at the early stage of COVID-19 in Wuhan, China (in blue) and Japan (in red).

**Figure 2:**
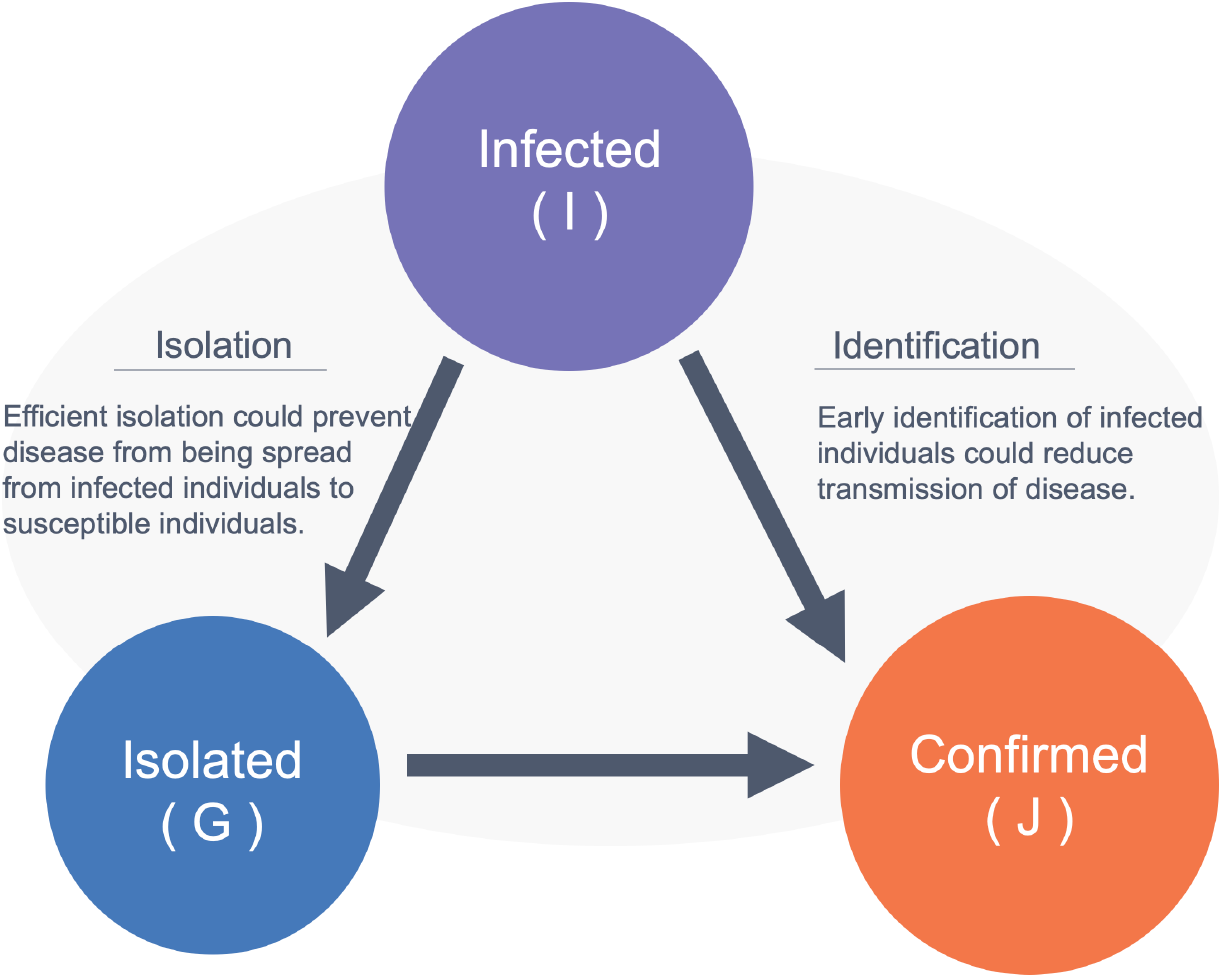
Sketch map of the model.

We make some interpretations for the model: from day *t* to *t* + 1,

1. The newly infected comes from a proportion (expressed as a growth rate *r*) of the potential infectious ones who might transmit the coronavirus to others.
2. The newly confirmed comes from the newly infected ones in history.
3. The infected ones are put into isolation (with isolation rate *ℓ*) once they show illness symptoms. And the newly confirmed should be removed from the isolated group.

### Quarantine strategy

We use a piecewise constant isolation rate *ℓ* to model the quarantine strategy:

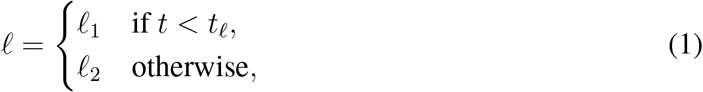

which means that at the day *t*_*ℓ*_, the quarantine strategy is changed to control the spread of the epidemic, leading to a change in the isolation rate.

### First infection date *t*_0_

Given the initial number of the infected *I*(*t*_0_), our model is able to track the first infection date *t*_0_ of the epidemic. It is REALLY important to find the first infection date statistically correct in different regions.

We fit the real data to identify the parameters, as listed in Table 1. Once the growth rate *r* is identified, we can determine the reproductive number *R*_0_ of COVID-19 according to (*8*).

**Table 1:** Parameters.

| | $r$ | $\ell_1$ | $\ell_2$ | $t_0$ | $t_\ell$ | $t_\ell - t_0$ |
| --- | --- | --- | --- | --- | --- | --- |
| Shanghai | 0.3137 | 0.1713 | 0.6149 | Dec.27 | Jan.16 | 20 |
| Beijing | 0.3125 | 0.1824 | 0.5880 | Dec.27 | Jan.17 | 21 |
| Hunan | 0.3098 | 0.0444 | 0.8421 | Dec.26 | Jan.19 | 24 |
| Zhejiang | 0.3129 | 0.1072 | 0.5950 | Dec.22 | Jan.15 | 24 |
| Henan | 0.3121 | 0.0592 | 0.5912 | Dec.25 | Jan.17 | 23 |
| Guangdong | 0.3118 | 0.1004 | 0.5679 | Dec.23 | Jan.16 | 24 |
| China without Hubei | 0.3072 | 0.1482 | 0.5973 | Dec.18 | Jan.18 | 31 |
| Hubei without Wuhan | 0.3135 | 0.1789 | 0.5084 | Dec.17 | Jan.17 | 31 |
| Wuhan | 0.3019 | 0.1142 | 0.4567 | Dec.17 | Jan.17 | 31 |

## COVID-19 in mainland China

In Figure 3, we plot the evolutions of *J/I* in several areas in mainland China. The x-axis represents the days started from the first infection date. From this figure we see that the curve for Beijing and Shanghai are ahead of Wuhan and Hubei, implying that earlier quarantine make the epidemic more controllable.

**Figure 3:**
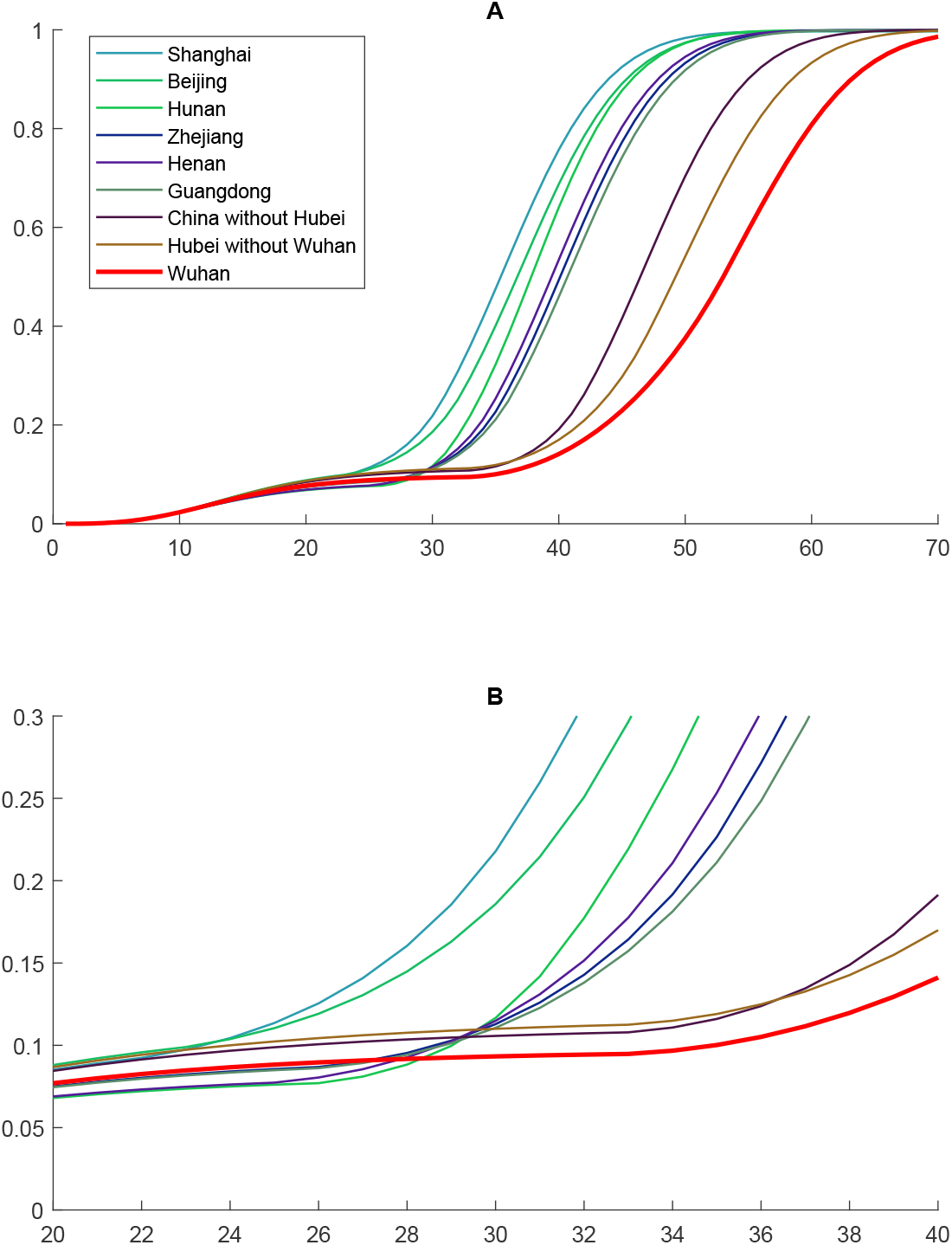
Fitting results of *J/I* for several areas in mainland China. (A) The whole time interval from the first infection date to the 70th day. (B) The local zoom from the 20th day to the 40th day since the first infection date *t*_0_ estimated by Fudan-CCDC model.

We show in Figure 4 the fitting results of the cumulative number of confirmed cases in several areas in mainland China. Data are presented in scattered red circles; the blue solid line is the fitting results of the data and its prediction, to which we applied Logistic regression 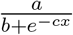 Our model is consistent with the published data for multiple areas outside Wuhan such as Shanghai. The discrepancy between the model and data for Wuhan could be explained by insufficient isolation and quarantine measures at the early stage of the epidemic. In other words, individuals with close contacts and suspicious exposure to confirmed cases might not have been isolated for a 14-day health observation period as later advised. Besides, the capacity of pathogenic diagnosis could not meet the need of viral nucleic acid test for suspected cases at the nascent stage. Therefore, our estimation of infected cases is higher than the published data.

**Figure 4:**
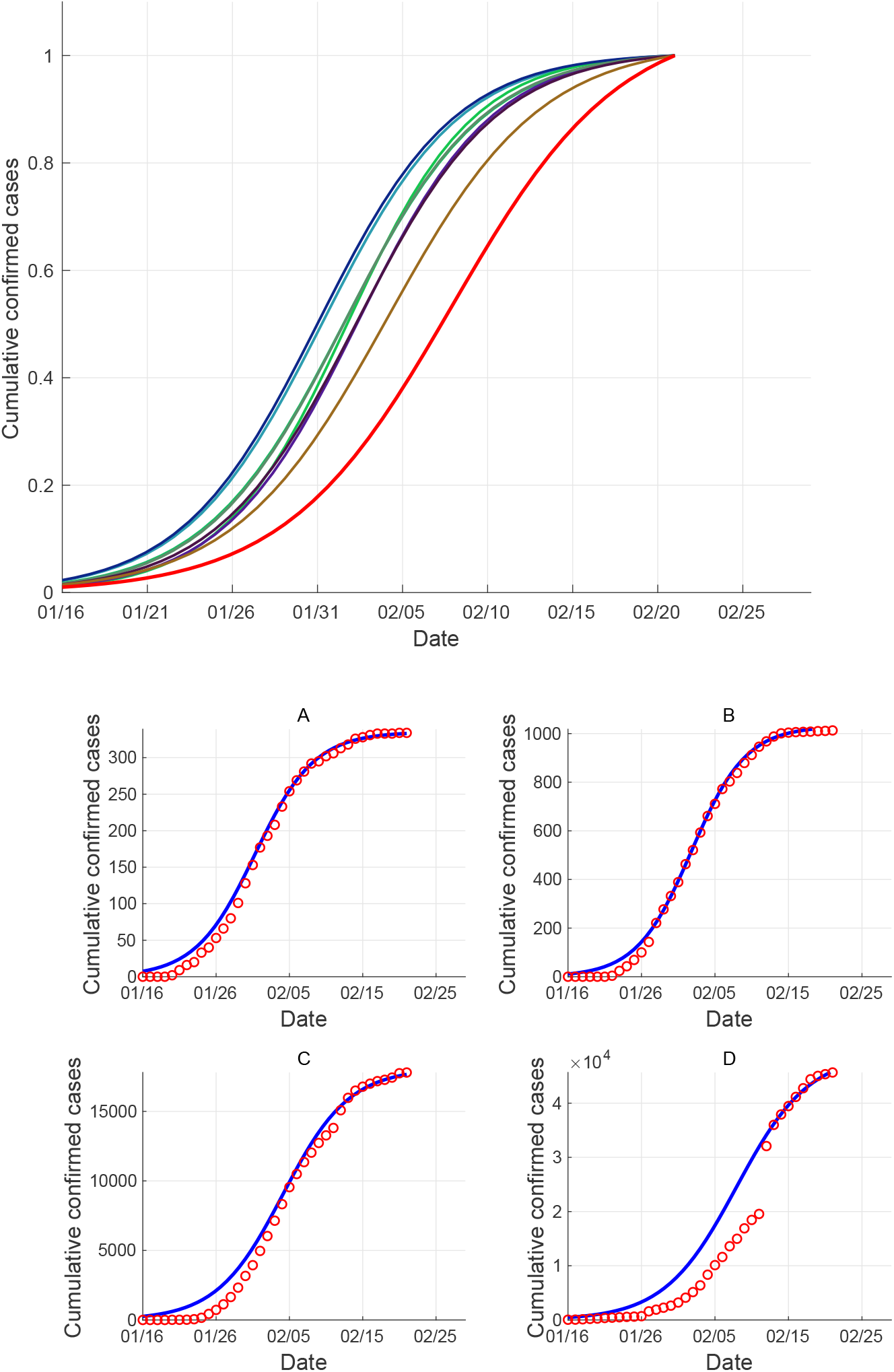
Top:Fitting results of the cumulative number of confirmed cases (normalized) in several areas in mainland China, legend same as in Figure 3; Bottom: Fitting results of the cumulative number of confirmed cases in (A) Shanghai, (B) Hunan, (C) Hubei Province (without Wuhan), (D) Wuhan.

We show in Figure 5 the fitting results of the daily confirmed cases in several areas in mainland China. The black line in the bottom subfigures represents the fitting results obtained from the derivatives of Logistic regressions.

**Figure 5:**
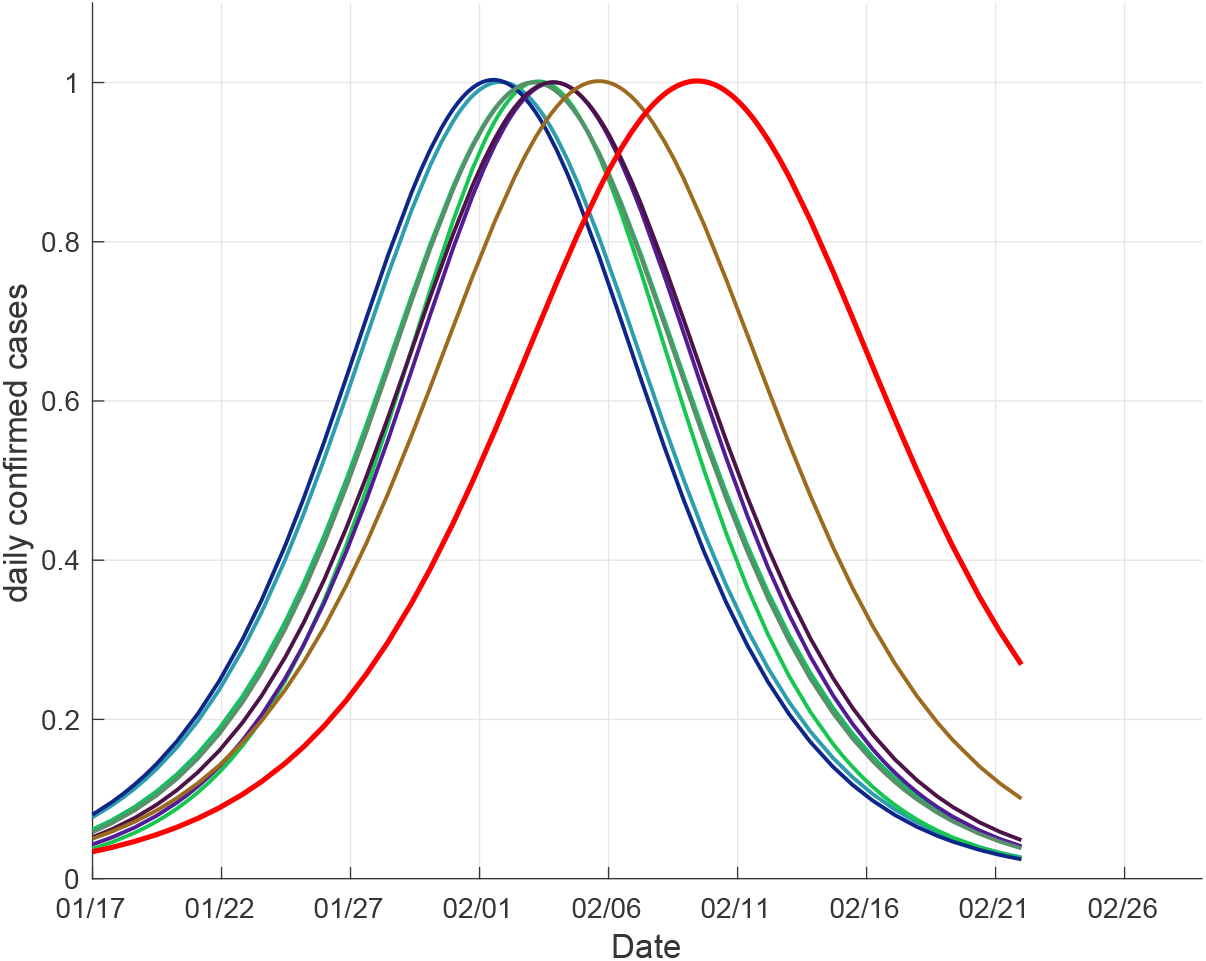
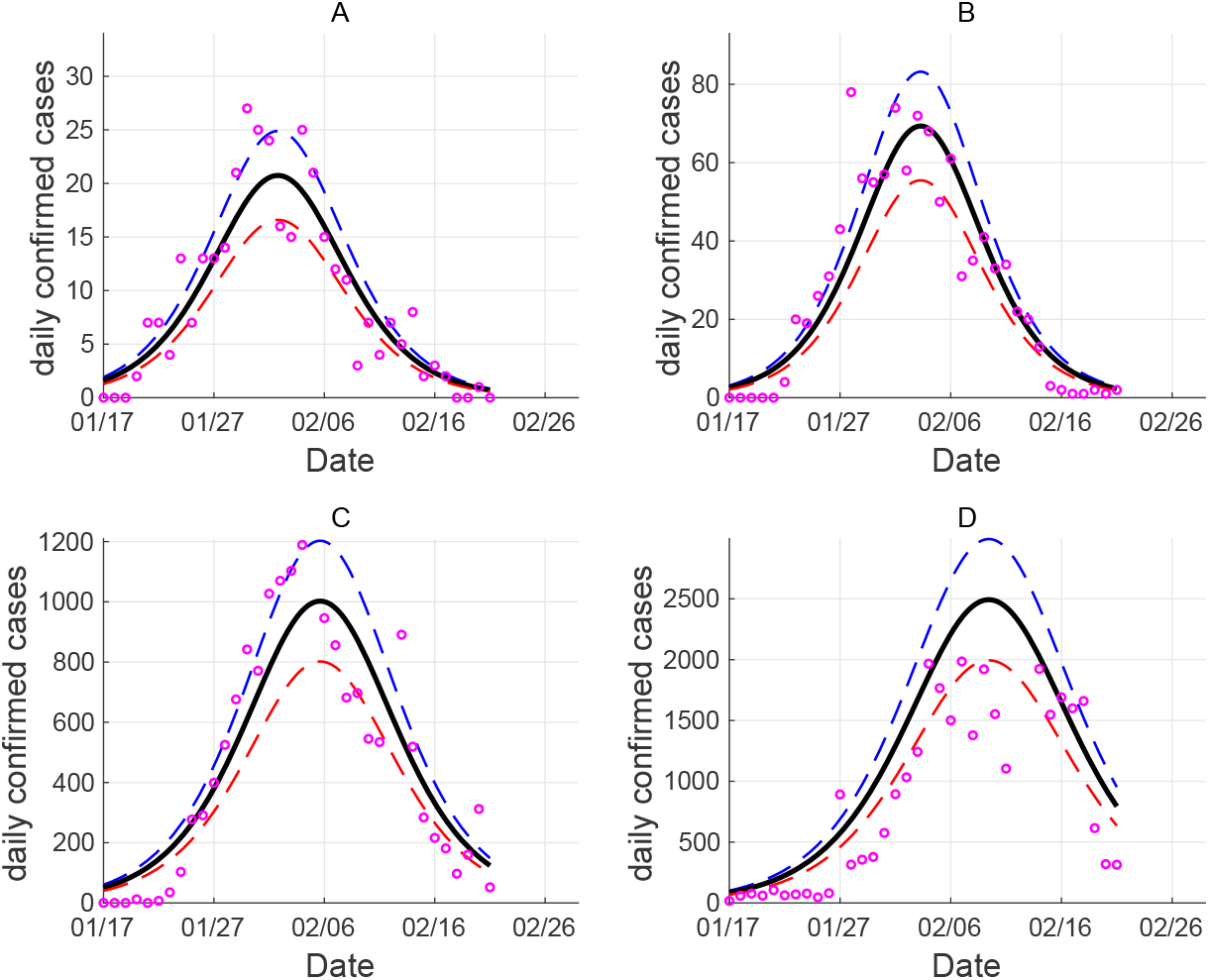
Top:Fitting results of the daily confirmed cases (normalized) in several areas in mainland China, legend same as in Figure 3; Bottom: Fitting results of the daily confirmed cases in (A) Shanghai, (B) Hunan Province, (C) Hubei Province (without Wuhan), (D) Wuhan.

We note that since no public data of the cumulative infected *I*(*t*) have been given, the number of the infected is always calculated by our model. As can be seen from the sub-figure A in Figure 6, if *ℓ*_2_ = *ℓ*_1_, the epidemic will continue to grow, while if *ℓ*_2_ *< ℓ*_1_, the epidemic in all nine areas will slow down. We can clearly see that the nine areas can be divided into three groups based on number of confirmed cases. For Beijing and Shanghai, they are the first to take effective quarantine measures, and receive the best control effects. The second group consists of Hunan, Zhejiang, Henan and Shangdong, of which the isolation rate is lower and thus have a fast confirm rate. Specially, in a later stage, Hunan started taking strong quarantine measures and received better control effects, compared to the rest three areas in this group. Lastly, the third group is made up of China (without Hubei), Hubei (without Wuhan) and Wuhan, which were the last to take quarantine measures. In particular, the average administrative strength of China (without Hubei) is stronger than that of Hubei (without Wuhan). Therefore, the average final control effects in China (without Hubei) is better than that of Hubei (without Wuhan).

**Figure 6:**
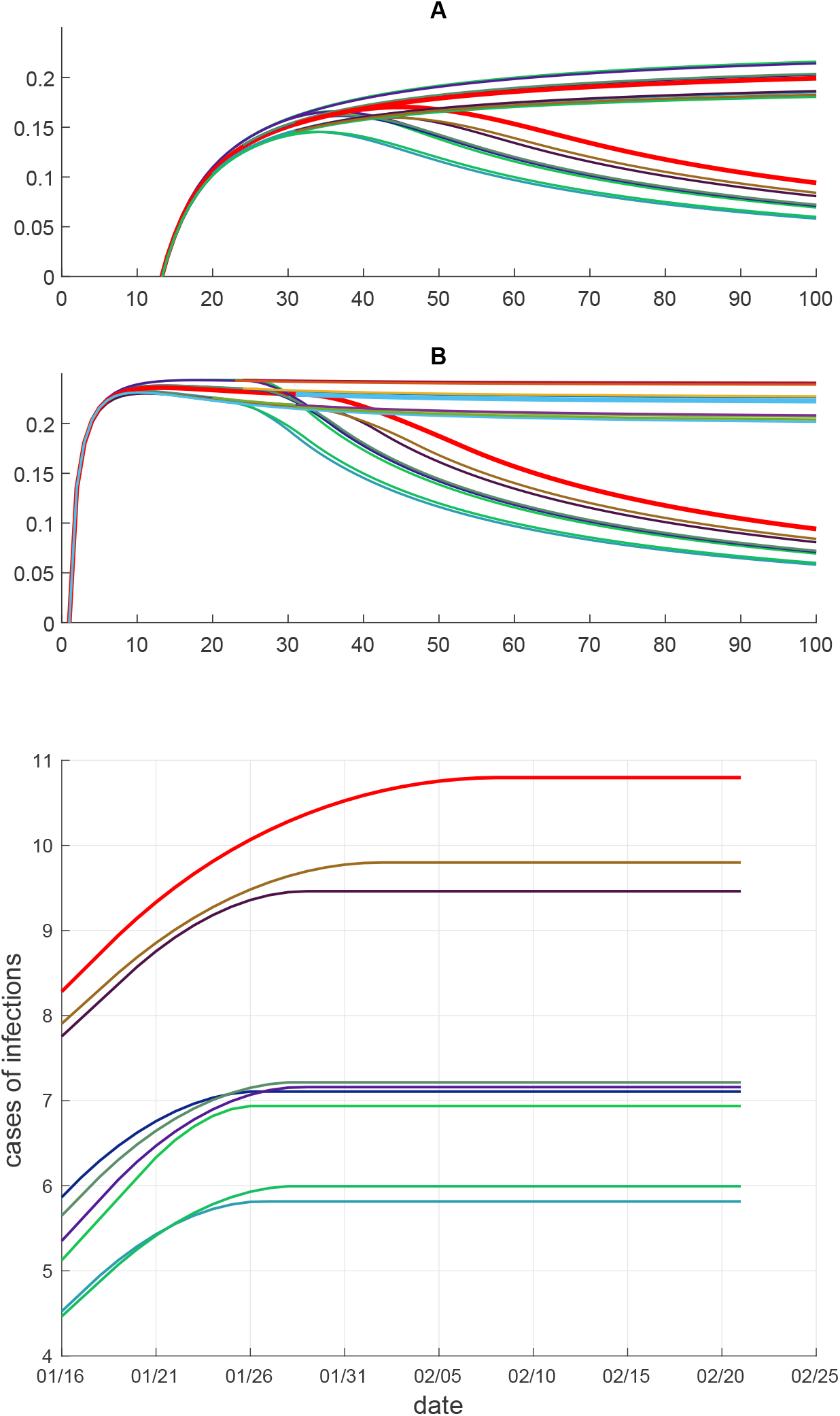
Top: Fitting results of the epidemic evolution in several areas in mainland China, with *ℓ*_2_ = *ℓ*_1_ and *ℓ*_2_ *ℓ l*_1_. The legend is the same as in Figure 3. (A) The evolution of 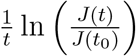 with respect to days started from the first infection date. (B) The evolution of 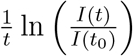 with respect to days started from the first infection date.Bottom: Fitting results of log of the cumulative infected number (log(*I*(*t*))) in several areas in mainland China.

In the sub-figure B in Figure 6, it can be seen that the trend of the infection number has a high similarity with that of the confirmed number. Areas with poorer early stage management, such as Hunan and Henan, have a faster disease spread rate. After the time all areas took effective quarantine measures, the curves evidently move downwards, which show that the epidemic is under control. Therefore, it’s crucial to take effective quarantine measures as early as possible, and to increase the isolation rate *ℓ*.

## Possible future scenarios for COVID-19 in Japan

In Figure 7, we present possible scenarios for epidemic evolution in Japan predicted by FUDAN-CCDC model. It can be clearly seen that the isolation rate *ℓ*_2_ can affect the containment of the epidemic. The final estimated number of infected people goes down as *ℓ*_2_ goes up. When *ℓ*_2_ = *ℓ*_1_ or *ℓ*_2_ = 0.35, namely when the quarantine strategy taken is not effective enough, the infected/confirmed number will grow exponentially. It can be observed that 0.40 seems a threshold for *ℓ*_2_, above which the infected number can be eventually under control, although when *ℓ*_2_ = 0.40 the final cumulative infection number is quite large. When *ℓ*_2_ = 0.6, the estimated scale of final cumulative infection is smaller, which is about 3000.

**Figure 7:**
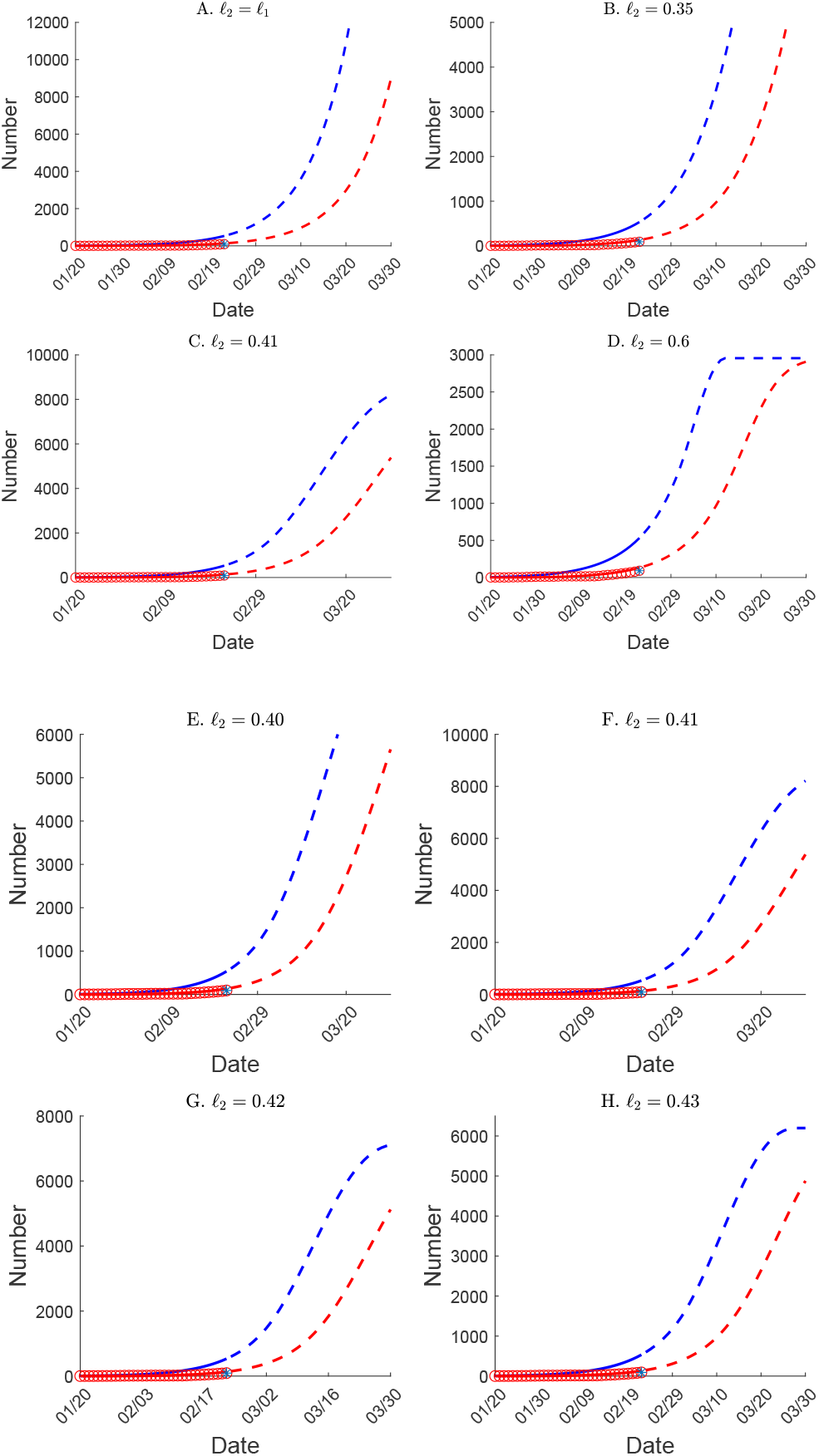
Fitting results of the epidemic evolution in Japan under four choices of *ℓ*_2_ started from March 1st, 2020. Blue: cumulative infected; Red: cumulative confirmed. (A) *ℓ*_2_ = *ℓ*_1_ = 0.3051. (B) *ℓ*_2_ = 0.35. (C) *ℓ*_2_ = 0.41. (D) *ℓ*_2_ = 0.6. (E) *ℓ*_2_ = *ℓ*_1_ = 0.40. (F) *ℓ*_2_ = 0.41. (G) *ℓ*_2_ = 0.42. (H) *ℓ*_2_ = 0.43.

We are worried about the current situation in Japan. We think that the quarantine strategy in Japan is not enough to fight the spread of COVID-19. We suggest Japan not to let the confirmed cases isolate at home. Instead, keeping all the confirmed cases at hospital is good for Japan.

## Discussion

Our findings suggest that effective isolation measures as early as possible are crucial for affected regions. **Based on our model, the future trend of the epidemic highly depends on when and how the measures will be adopted**. Effective interventions include limiting population mobility (e.g. cancellation of mass gathering, school closures and work-from-home arrangements) and public education (e.g. use of face masks and improved personal hygiene) (*1*). Given that the effects of public health interventions will usually lag by a period of time, it is important to make adjustment on corresponding measures as soon as the epidemic changes. In order to establish effective measures, the data of confirmed and suspected cases should be as accurate as possible. The accuracy depends on the potency of diagnosis. Guidelines on diagnosis and therapy need to fit current situation. For example, on Feb 12, 2020, National Health Commission of China recommended to make clinical diagnosis besides pathogenic diagnosis when there were not enough kits and facilities to perform viral nucleic acid tests. We hope that suggestions from Chinese doctors and researchers could be seriously considered worldwide and that the situation in Wuhan could be discussed carefully. Just as the saying goes, things are always like this: COVID-19 is not likely to come once everything is in a state of preparedness, whereas an outbreak could hardly be avoided.

## Data Availability

The data from Wind database or any one can find them in the website of Chinese Center for Disease Control and Prevention(CCDC). Only the cumulative numbers in hospitalization are needed.

## Acknowledgments

We should truly thank Prof. Tatsien Li. We also thank Yongzhen Wang, Yun Wang, Prof. Rongmin Li and Prof. Shanjian Tang(Fudan), Gang Liang(China CDC in Wuhai, Hubei), Dr. Zhihua Shen(Fusion Fin Trade, fusionfintrade.com),, Ling Ye(China CDC in Daishan, Zhejiang), Long Chen(University of California, Irvine), Prof. Jiongmin Yong(University of Central Florida), Prof. Zhaojun Bai(UC Davis) and Xinkang Cao(Mathworks). Wenbin Chen thanks Ning Liu in the Winning Health Technology Group Company(winning.com.cn), whose group always believe and support us. Wenbin Chen also thanks his wife Dr. Jie Xu, whose hometown is Wuhan.

We should thanks everyone who are fighting for Wuhan, specially we present our deep respects to Doctor Wenliang Li and Prof. Nanshan Zhong.

## Authors contributions

The simulations are main implemented by Shao Nian and designed by Wenbin Chen. All authors conceived the study, carried out the analysis, discussed the results, drafted the first manuscript, critically read and revised the manuscript, and gave final approval for publication.

## Confiict of interests

The authors declare no competing interests.

## Data and materials availability

The data employed in this paper are acquired from the National Health Commission of China (http://www.nhc.gov.cn), and Ministry of Health, Labour, and Welfare, Japan (https://www.mhlw.go.jp/index.html). All the data can be accessed publicly. No other data are used in this paper.

